# Asymmetrical Frontal–Subcortical Aberrance in ADHD: Empirical Support of a Developmental Network Model

**DOI:** 10.1101/2025.06.25.25330293

**Authors:** Tien-Wen Lee

## Abstract

**Background:** This study empirically tests a theoretical model of ADHD that identifies aberrant frontal-to-subcortical projections—particularly those involving the ventral tegmental area and, consequently, the nucleus accumbens (NAc)—as a core neurobiological vulnerability. The model predicts reduced frontal influence on subcortical structures and group-by-direction interaction effects, especially at the NAc.

**Methods:** Structural and resting-state fMRI data from sixty individuals with ADHD and sixty healthy controls were selected from the ADHD-200 dataset. The cortex was parcellated using MOSI (Modular Analysis and Similarity Measurements). The mean amplitude of low-frequency fluctuations (ALFF) and the mean time series were extracted for each frontal node. Subcortical nuclei—including the caudate, putamen, globus pallidus, NAc, and thalamus—were included in the analyses, with each voxel treated as a neural node. Correlation analyses were performed to examine the relationship between nodal power (ALFF) and nodal degree. Directional metrics were defined as frontal-to-subcortical (F2S) and subcortical-to-frontal (S2F) correlations between nodal strength and nodal power. Group differences and interactions were tested using t-tests and linear mixed-effects (LME) models.

**Results:** Correlation and LME analyses revealed a consistent bimodal pattern, with positive F2S and negative S2F associations across all subcortical nuclei, suggesting a directional excitation–inhibition balance. Reduced F2S slopes in ADHD and a significant group-by-direction interaction in the NAc (p = 0.009) supported model predictions.

**Conclusions:** The proposed developmental model of ADHD was empirically supported.

## Introduction

Attention-deficit/hyperactivity disorder (ADHD) ranks among the most common neurodevelopmental conditions, with an estimated prevalence of approximately 6% in children and adolescents and 2.5% in adults (Willcutt, 2012, American Psychiatric Association, 2013). Clinically, ADHD is categorized into three primary subtypes: predominantly inattentive, predominantly hyperactive/impulsive, and combined presentation. In addition to the core symptoms, affected individuals commonly exhibit a broader range of neurocognitive and emotional disturbances. These may include executive dysfunction, emotion dysregulation, difficulties with planning and organization, impaired impulse control, problems in social interaction, hypoarousal, heightened response time variability, and altered reward sensitivity, which together shape the broader clinical picture (Barkley et al., 2001, Fuermaier et al., 2017, Kofler et al., 2013, Schweitzer and Sulzer-Azaroff, 1995, Taş Torun et al., 2022, Du Rietz et al., 2019, Souroulla et al., 2019, Boucsein, 2012, Devilbiss and Waterhouse, 2011, An et al., 2013). A wide spectrum of psychiatric and neurological comorbidities has also been documented, including tic disorders, anxiety, depression, bipolar disorder, autism spectrum disorder, and, in some cases, an increased risk of Parkinson’s disease later in life (Schiweck et al., 2021, McIntosh et al., 2009, Hours et al., 2022, Radomyslsky et al., 2025). Although the formal diagnostic threshold requires symptom onset before age 12, early behavioral signs—such as excessive motor activity, poor sustained attention, and impulsivity—can often be detected during the preschool years, typically between ages 2 and 5. Rather than emerging abruptly, these symptoms usually unfold gradually over the course of development (Egger and Angold, 2006).

Consistent with its heterogeneous clinical manifestations, ADHD is associated with widespread neural abnormalities spanning both the dorsal and ventral fronto-striatal circuits, limbic structures, and deep brain nuclei—most notably the ventral tegmental area (VTA) and locus coeruleus (LC), which serve as central hubs for dopaminergic and noradrenergic signaling, respectively (Ernst et al., 1998, Ernst et al., 1999, Schneider et al., 2010, Cubillo et al., 2011, Rubia et al., 2009, Brieber et al., 2007, Ohno, 2003, Lukito et al., 2020, Norman et al., 2016, Frodl and Skokauskas, 2012, Connaughton et al., 2022, Bralten et al., 2016, Cortese et al., 2012, Depue et al., 2010, Castellanos et al., 2002, von Rhein et al., 2015, van Dongen et al., 2015, Strohle et al., 2008, Costa Dias et al., 2013). While complexity is intrinsic to nearly all psychiatric conditions, dialectic neuroscience—an approach grounded in centralized dialectics— offers a framework for developing integrative theories of psychopathology (Lee, 2025c). This approach has been applied to major depressive disorder, bipolar disorder, and autism spectrum disorder (Lee and Xue, 2017, Lee and Xue, 2018), with theoretical models subsequently supported by empirical findings (Lee, 2025b, Lee, 2025e, Lee, 2025d). In the case of ADHD, a candidate theory has recently emerged, positing that aberrant frontal-to-subcortical projections during development constitute a core neurobiological vulnerability (briefly outlined below), offering a unifying framework to reconcile a wide range of seemingly disparate or conflicting findings (Lee, 2025a). The present study aims to empirically evaluate this theoretical account.

Aberrant frontal-to-subcortical projections may initiate a cascade of network-level dysregulations, with the VTA and nucleus accumbens (NAc) highlighted as central foci of neuropathological disruption in ADHD. Hypoactive glutamatergic signaling from the frontal cortex (FCX) to the VTA may suppress tonic dopamine (DA) levels throughout mesocorticolimbic circuits, compromise dopaminergic feedback to the FCX, and aggravate hypofrontality (Floresco et al., 2001, Polter and Kauer, 2014). Attenuated frontal input may also lead to sensitization and disinhibition of the NAc (Floresco et al., 2003, Goto and Grace, 2005). In parallel, reduced tonic DA release from the VTA may enhance phasic dopaminergic responsivity reflected in the NAc (Grace, 1991, Grace, 1995), collectively contributing to aberrant reward processing and limbic hyperactivity. The latter may further accentuate hypofrontality via reciprocal inhibitory mechanisms (Lee and Xue, 2018, Lee, 2025b). A similar dysregulation may affect the dorsal fronto-striatal pathway, as reflected in structural and functional abnormalities in the caudate and putamen (Brieber et al., 2007, Ohno, 2003, Lukito et al., 2020, Norman et al., 2016, Connaughton et al., 2022). To empirically test the model, it is essential to establish directional inferences—both frontal-to-subcortical (F2S) and subcortical-to-frontal (S2F), with specific emphasis on the NAc. Two predictions were proposed in the previous article (Lee, 2025a): first, the influence from the FCX to subcortical nuclei is weakened in ADHD; second, a group-by-direction interaction should be observable in directional neural metrics.

Resting-state functional magnetic resonance imaging (rsfMRI) data from the public ADHD200 dataset will be used to test the proposed hypotheses (Consortium, 2012), where causality analysis is essential for this purpose. While correlation is inherently unidirectional, recent research suggests that in cross-modal contexts, certain causal inferences are still feasible (Lee and Tramontano, 2025). This approach has been employed in major depressive disorder to assess the hypothesis of compartment-level mutual suppression (Lee, 2025b). Specifically, nodal strength (i.e., the total functional connectivity of a node) appears to influence nodal power (i.e., ALFF), but not vice versa. Following prior studies, two directional metrics were used to characterize the relationship between nodal strength and nodal power within the frontal–subcortical network. Subcortical nuclei were incorporated based on the canonical fronto-striato-thalamo-cortical loops (Alexander and Crutcher, 1990, Grahn et al., 2008), which encompass the caudate (Cau), nucleus accumbens (NAc), globus pallidus (Pal), putamen (Put), and thalamus (Tha), see Figure 1. The “F2S” index refers to the association between frontal-to-subcortical nodal strength and subcortical ALFF, whereas “S2F” captures the reverse—subcortical-to-frontal nodal strength versus frontal ALFF. Functional parcellation of the cortex will be performed using MOSI (Modular Analysis and Similarity Measurements) (Lee and Tramontano, 2021). The core neuropathology of ADHD is expected to emerge through a two-way interaction effect, reflecting slope differences between F2S and S2F across ADHD and healthy control (HC) groups (ADHD vs. HC × FCX-to-NAc vs. NAc-to-FCX).

**Figure 1.**
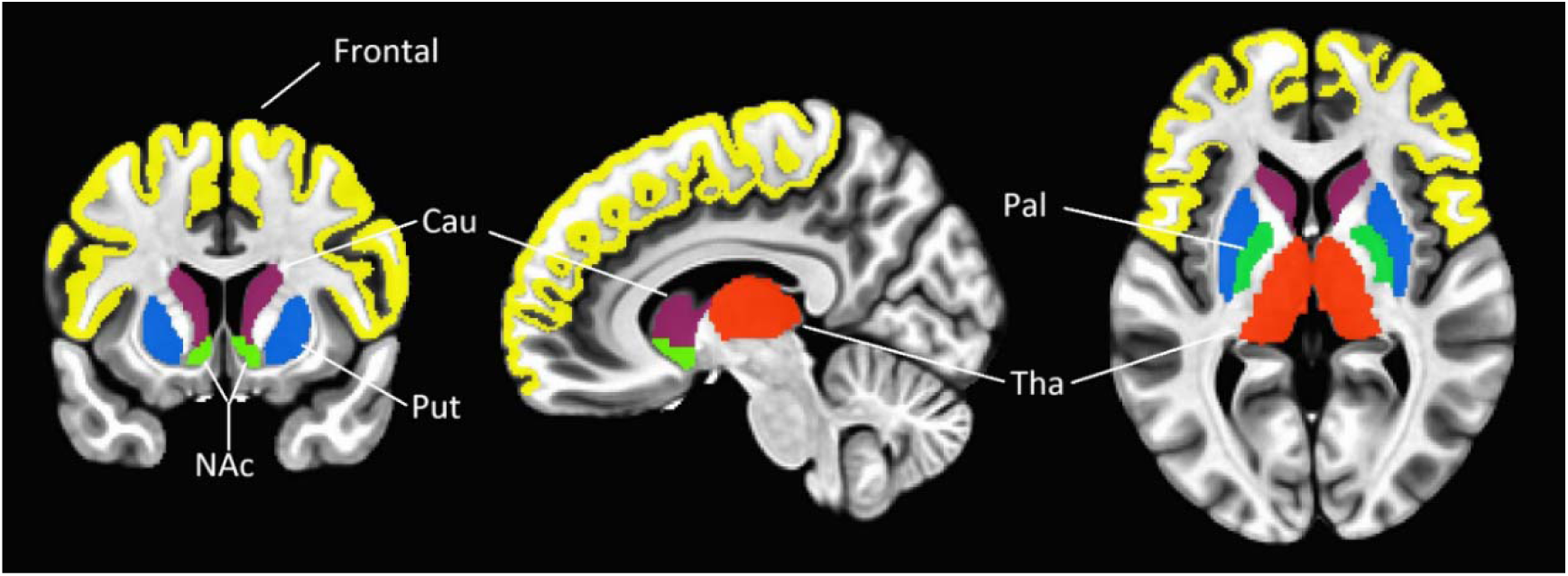
An illustration of the frontal lobe and subcortical nuclei—Cau (caudate), NAc (nucleus accumbens), Pal (globus pallidus), Put (putamen), and Tha (thalamus), each highlighted in a distinct color.

## Materials and Methods

### MRI datasets and preprocessing

We employed data from 120 participants (60 with ADHD and 60 healthy controls) obtained from the ADHD-200 dataset (New York University Child Study Center) (Bellec et al., 2017, Consortium, 2012), with written informed consent provided by all individuals. The functional and structural MRI images of the entire brain were recorded using 3.0 Tesla Siemens MRI machine (fMRI: TR=2s, TE=15ms, Flip Angle=90°, voxel size=3×3×4mm%3, 33 slices; 170 volumes).

rsfMRI preprocessing was performed using AFNI (Cox, 1996). Steps included despiking, slice timing correction, motion realignment, coregistration to structural T1 images, spatial smoothing with a 6 mm FWHM kernel, and temporal bandpass filtering (0.01–0.1 Hz) via the 1dBport function, which generates sinusoidal regressors for frequency isolation. The first four volumes were discarded to ensure magnetization stabilization. T1-weighted anatomical images were processed with FreeSurfer to segment gray and white matter and to parcellate the cortex into 68 regions (34 per hemisphere) based on the Desikan-Killiany atlas (https://surfer.nmr.mgh.harvard.edu/fswiki/CorticalParcellation) (Dale et al., 1999, Fischl et al., 1999, Desikan et al., 2006). These anatomical parcels served as seeds for subsequent MOSI-based parcellation. In line with previous pipelines (Lee et al., 2014, Xue et al., 2014), nuisance variables—including 12 motion parameters, white matter, and CSF signals—were regressed out. A second-order polynomial was also fitted to model baseline drift. ALFF was calculated as the square root of signal power within the bandpass-filtered EPI data.

### ALFF, Functional connectivity and MOSI analysis

ALFF reflects spontaneous neural activity by quantifying the amplitude of low-frequency fluctuations in the resting-state BOLD signal, computed as the square root of power within the band-pass filtered spectrum. Due to the small size and internal heterogeneity of subcortical nuclei, along with the absence of standardized partition in rsfMRI studies, no further parcellation was applied to these subcortical regions. Consequently, MOSI-based parcellation was restricted to the cortical areas. In other words, the neural node is defined as a voxel within subcortical nuclei, whereas in the FCX, a neural node corresponds to a MOSI-partitioned module (cluster). Functional connectivity (FC) was measured via Fisher-transformed Pearson correlation coefficients, calculated between voxel-wise time series in subcortical areas and cluster-averaged time series in the FCX. Mean nodal strength was defined as the total FC centered on each subcortical voxel or frontal module.

The FC-derived connectivity matrix served as the input for MOSI, a cortical parcellation framework combining modular decomposition and similarity-based aggregation (Lee and Tramontano, 2021). The method iteratively partitions the cortex by first identifying local functional modules via community detection—implemented using the Louvain algorithm (Blondel et al., 2008, Reichardt and Bornholdt, 2006)—and subsequently merging spatially contiguous modules exhibiting high functional similarity. This procedure continues until the parcellation stabilizes. In summary, MOSI is governed by two principles: spatial adjacency and functional coherence, ensuring that neighboring voxels with similar time series are grouped together—an essential premise of functional parcellation. Voxels forming isolated modules of fewer than five were discarded to minimize the influence of spurious noise. The granularity of the final parcellation is adjustable via the resolution parameter gamma, which controls the number of resultant modules. Higher gamma values yield finer partitions. MOSI’s performance was evaluated through intra-module homogeneity and benchmarked against standard atlas-based schemes (see **Results**).

While the full MOSI procedure was initially conducted across a broad gamma range (0.40 to 0.85), consistent with prior work involving whole-cortex partitioning (note: only the FCX was examined in this study) (Lee, 2025b), further constraints were required for the formal analysis. Given that correlation analyses require a minimum of 30 data points for stable and interpretable estimates (Bonett and Wright, 2000), the selection of modular resolution was accordingly limited. Empirically, a gamma value of 0.70 yielded an average of approximately 38 modules within the FCX for both ADHD and control groups. Therefore, only gamma values ≥ 0.70 (i.e., 0.70, 0.75, 0.80, 0.85) were included in the main analysis to ensure adequate statistical power for correlation-based measures.

### Formal analysis I—correlation analyses between ALFF and nodal strength

Two types of correlation analyses were conducted to examine the association between ALFF and nodal strength. The first focused on fronto-to-subcortical connectivity, correlating nodal strength from frontal modules with ALFF within subcortical regions, including Cau, NAc, Pal, Put, Tha, and the combined voxel set encompassing all five nuclei (F2S). To account for variability in module size across the FCX, a size-weighted averaging strategy was applied when computing representative (mean) nodal strength. The second analysis reversed the direction, assessing subcortico-to-frontal connectivity. Here, mean ALFF was calculated for each frontal module, and mean nodal strength was computed voxel-wise within the subcortical nuclei (S2F). As the subcortical unit of analysis was defined at the voxel level, no additional weighting was required.

### Formal analysis II—linear mixed-effects modeling of ALFF and nodal strength

The author employed a linear mixed-effects model (LME) to investigate how diagnostic group (*Group*: ADHD vs. healthy control [HC]), brain region (*Region*: frontal to subcortical nucleus vs. subcortical to frontal), and mean nodal strength (*d*) contribute to the prediction of mean ALFF (*a*), the dependent variable. Both ALFF and mean nodal strength were normalized prior to inclusion in the model. The variable *Subject* indexed individual participant, each contributing repeated measurements across both regions. The model included fixed effects for Group, Region, *d*, and all interactions among them, including the three-way interaction (Group × Region × *d*). A random intercept was included for each Subject to account for within-subject dependencies due to repeated measures. The model was expressed as: ***a = β₀ + β₁·d + β₂·Group + β₃·Region + β₄·(d × Group) + β₅·(d × Region) + β₆·(Group × Region) + β₇·(d × Group × Region) + u₀[Subject] + ε***, where *u₀[Subject]* represents the subject-specific random intercept, and *ε* is the residual error. This model allows each participant to have their own baseline level of ALFF, while estimating group-level and interaction effects assuming a common slope for nodal strength across subjects. The analysis was implemented using the fitlme function in MATLAB R2020a (The MathWorks, Natick, MA, USA), with the model specification: ***a ∼ Group * Region * d + (1 | Subject)***.

An alternative, more complex model was considered, which included random slopes for the continuous predictor b at the subject level, specified in MATLAB as: ***a ∼ Group*Region*d + (1+d|Subject)***. This model allows individual subjects to have varying slopes with respect to b, capturing subject-specific differences in the effect of b on the outcome. However, model comparison based on Akaike Information Criterion (AIC), Bayesian Information Criterion (BIC), and log-likelihood favored the simpler model. The simpler model exhibited consistently lower AIC and BIC values and a higher log-likelihood, indicating better overall fit and parsimony (detailed results are presented in Part III of the **Supplementary Materials**). Given the nature of the data—smoothed functional MRI measurements centered on small subcortical structures, where spatial smoothing reduces local variability—the inclusion of random slopes did not provide meaningful improvement and posed a risk of overfitting. Consequently, the simpler random intercept model was adopted as the primary analytical framework.

For the LME analyses, as the results across different gamma values are not statistically independent, a representative p-value was computed using the geometric mean. Specifically, for four gamma values yielding p₁, p₂, p₃, and p₄, the representative p-value was calculated as (p₁ × p₂ × p₃ × p₄)^(1/4). A Bonferroni-corrected threshold of p < 0.01 (i.e., 0.05/5 for the five subcortical nuclei) was applied for statistical significance.

## Results

Demographic characteristics were comparable between the combined type ADHD and HC groups (n = 60 each), as sourced from the New York University Child Study Center dataset of the ADHD-200 Consortium (https://fcon_1000.projects.nitrc.org/indi/adhd200/). There was no significant difference in age between groups (ADHD: 10.1 ± 1.4 years; HC: 10.1 ± 1.7 years; *t*(118) = 0.0046, *p* = 0.9964). Gender distribution also did not differ significantly: ADHD group had 42 males and 18 females, while the HC group had 33 males and 27 females. A chi-square test with Yates’ correction yielded χ² = 2.28, *p* = 0.1314.

Group comparisons of subcortical volumes between children with ADHD and healthy controls (HC) revealed no statistically significant differences across the five examined nuclei (see Part I of the **Supplementary Materials** for details). Similarly, no significant between-group difference was observed in frontal lobe volume, with the ADHD group showing 6217.4 ± 543.9 voxels and the HC group 6421.8 ± 679.9 voxels (p = 0.071, t = -1.82). The trend toward reduced frontal volume in the ADHD group aligns with previous findings in the literature (Shaw et al., 2007, Norman et al., 2016). The effectiveness of MOSI-based functional cortical parcellation is illustrated in Figure 2.

**Figure 2.**
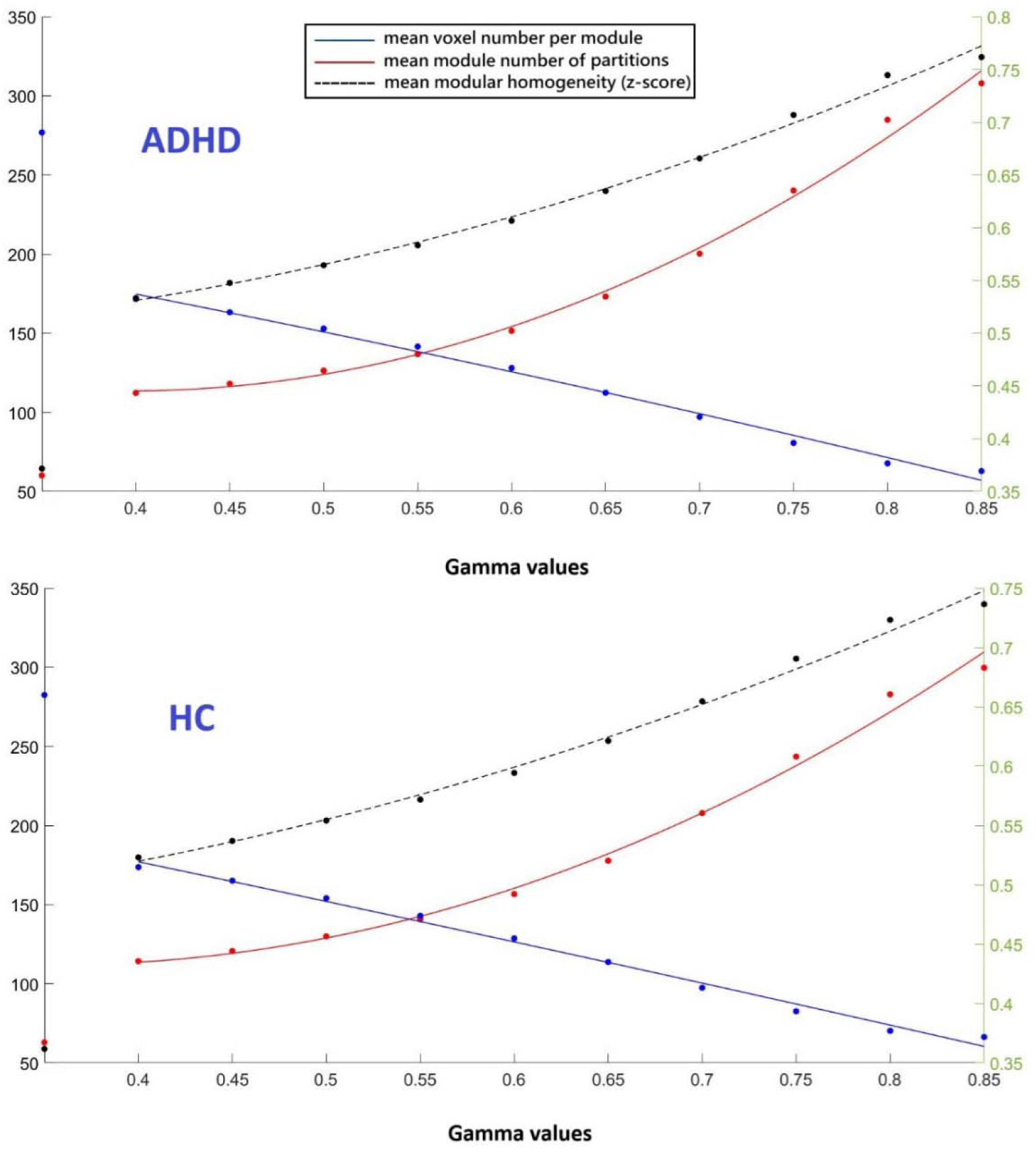
Scatter plots showing three metrics derived from the ADHD200 dataset across 10 gamma values (X-axis), with second-order polynomial regression curves overlaid. Each plot contains 10 data points per metric, corresponding to the tested gamma values. The top panel illustrates results for the ADHD group, while the bottom panel shows those for the HC group. The point at x = 0 denotes the initial parcellation using the Desikan-Killiany atlas. Red markers and curves represent the mean number of modules (left Y-axis), blue markers and curves indicate the average voxel count per module (left Y-axis), and black markers and curves denote the mean within-module z-score (right Y-axis), reflecting modular homogeneity.

### Analysis I—correlation analyses between ALFF and nodal strength

This study is the first to examine the association between nodal strength and nodal power within the fronto-subcortical network. While prior studies applying similar analytic approaches to cortical regions have reported a negative association— interpreted as reflecting a net inhibitory effect, whereby higher nodal strength corresponds to lower ALFF—the present findings in the fronto-striatal system reveal a distinct bimodal pattern. Specifically, F2S connections consistently exhibited a positive association with ALFF, whereas S2F connections showed a negative association, observed across both groups and robust across all gamma thresholds. This directional dissociation—characterized by uniformly positive F2S–ALFF and negative S2F–ALFF estimates across subcortical nuclei—is evident in both the correlation analyses (Tables 1 and 2) and the slope estimates derived from LME models (Table 3 in the main text; Table S4 in the Supplementary Materials). Together, these findings suggest a dynamic interplay of excitatory and inhibitory influences underlying fronto-subcortical communication. In the HC group, most correlations reached statistical significance, except for a positive trend in NAc under the F2S condition and negative trends in Cau and NAc under the S2F condition. In the ADHD group, a positive trend was observed in NAc under the F2S condition, with the remaining correlations showing statistical significance.

**Table 1.** Correlation between subcortical ALFF and fronto-subcortical connectivity (F2S)

|  | ADHD |  | one-sample t-tests |  | HC | one-sample t-tests |  |
| --- | --- | --- | --- | --- | --- | --- | --- |
| Gamma | mean (std) | t-scores | p-values |  | mean (std) | t-scores | p-values |
| **Cau** |  |  |  |  |  |  |  |
| 0.70 | 0.13 (0.26) | 3.74 | 4.14E-04 |  | 0.15 (0.20) | 5.85 | 2.31E-07 |
| 0.75 | 0.13 (0.26) | 3.85 | 2.97E-04 |  | 0.15 (0.20) | 5.93 | 1.70E-07 |
| 0.80 | 0.13 (0.26) | 3.86 | 2.86E-04 |  | 0.15 (0.20) | 5.94 | 1.61E-07 |
| 0.85 | 0.12 (0.26) | 3.67 | 5.22E-04 |  | 0.16 (0.20) | 6.09 | 9.26E-08 |
| **NAc** |  |  |  |  |  |  |  |
| 0.70 | 0.09 (0.36) | 2.02 | 4.82E-02 |  | 0.09 (0.32) | 2.07 | 4.28E-02 |
| 0.75 | 0.09 (0.35) | 2.05 | 4.53E-02 |  | 0.08 (0.31) | 1.88 | 6.45E-02 |
| 0.80 | 0.09 (0.36) | 1.86 | 6.81E-02 |  | 0.06 (0.32) | 1.55 | 1.26E-01 |
| 0.85 | 0.09 (0.36) | 1.93 | 5.83E-02 |  | 0.05 (0.32) | 1.15 | 2.54E-01 |
| **Pal** |  |  |  |  |  |  |  |
| 0.70 | 0.10 (0.23) | 3.33 | 1.50E-03 |  | 0.13 (0.23) | 4.29 | 6.63E-05 |
| 0.75 | 0.10 (0.23) | 3.38 | 1.28E-03 |  | 0.13 (0.23) | 4.45 | 3.85E-05 |
| 0.80 | 0.10 (0.22) | 3.68 | 5.01E-04 |  | 0.13 (0.22) | 4.52 | 2.99E-05 |
| 0.85 | 0.10 (0.23) | 3.46 | 1.00E-03 |  | 0.13 (0.22) | 4.47 | 3.65E-05 |
| **Put** |  |  |  |  |  |  |  |
| 0.70 | 0.13 (0.20) | 5.10 | 3.75E-06 |  | 0.11 (0.19) | 4.68 | 1.74E-05 |
| 0.75 | 0.13 (0.20) | 5.04 | 4.63E-06 |  | 0.12 (0.19) | 4.98 | 5.88E-06 |
| 0.80 | 0.12 (0.19) | 4.93 | 7.02E-06 |  | 0.11 (0.19) | 4.50 | 3.29E-05 |
| 0.85 | 0.13 (0.19) | 5.17 | 2.88E-06 |  | 0.11 (0.19) | 4.73 | 1.45E-05 |
| **Tha** |  |  |  |  |  |  |  |
| 0.70 | 0.09 (0.23) | 3.20 | 2.21E-03 |  | 0.11 (0.20) | 4.33 | 5.76E-05 |
| 0.75 | 0.10 (0.22) | 3.29 | 1.70E-03 |  | 0.12 (0.20) | 4.48 | 3.47E-05 |
| 0.80 | 0.10 (0.22) | 3.37 | 1.32E-03 |  | 0.12 (0.19) | 4.61 | 2.18E-05 |
| 0.85 | 0.09 (0.23) | 3.12 | 2.80E-03 |  | 0.12 (0.20) | 4.63 | 2.06E-05 |
| **Subcortical Nuclei** |  |  |  |  |  |  |  |
| 0.70 | 0.11 (0.18) | 4.40 | 4.53E-05 |  | 0.11 (0.15) | 5.83 | 2.50E-07 |
| 0.75 | 0.11 (0.18) | 4.54 | 2.85E-05 |  | 0.12 (0.14) | 6.25 | 5.05E-08 |
| 0.80 | 0.11 (0.18) | 4.55 | 2.69E-05 |  | 0.11 (0.14) | 6.17 | 6.62E-08 |
| 0.85 | 0.10 (0.18) | 4.38 | 4.92E-05 |  | 0.12 (0.15) | 6.16 | 7.10E-08 |

2 N = 60. Correlation coefficients were Fisher z-transformed. One-sample t-tests against zero were 3 conducted, with p-values reported in scientific notation and thresholded at 0.01. **Subcortical Nuclei** 4 indicates lumping all 5 nuclei together. All one-sample t-tests yielded positive t-values, whereas all two- 5 sample t-tests were statistically non-significant.

**Table 2.**
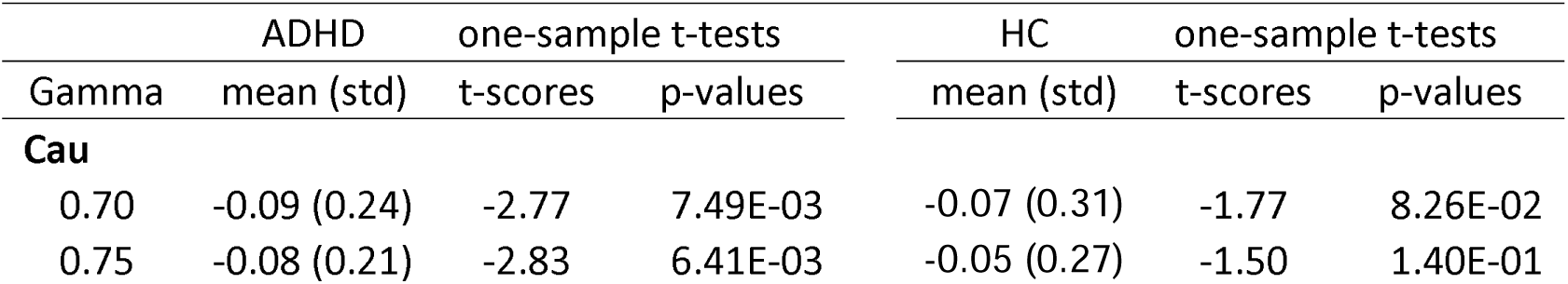
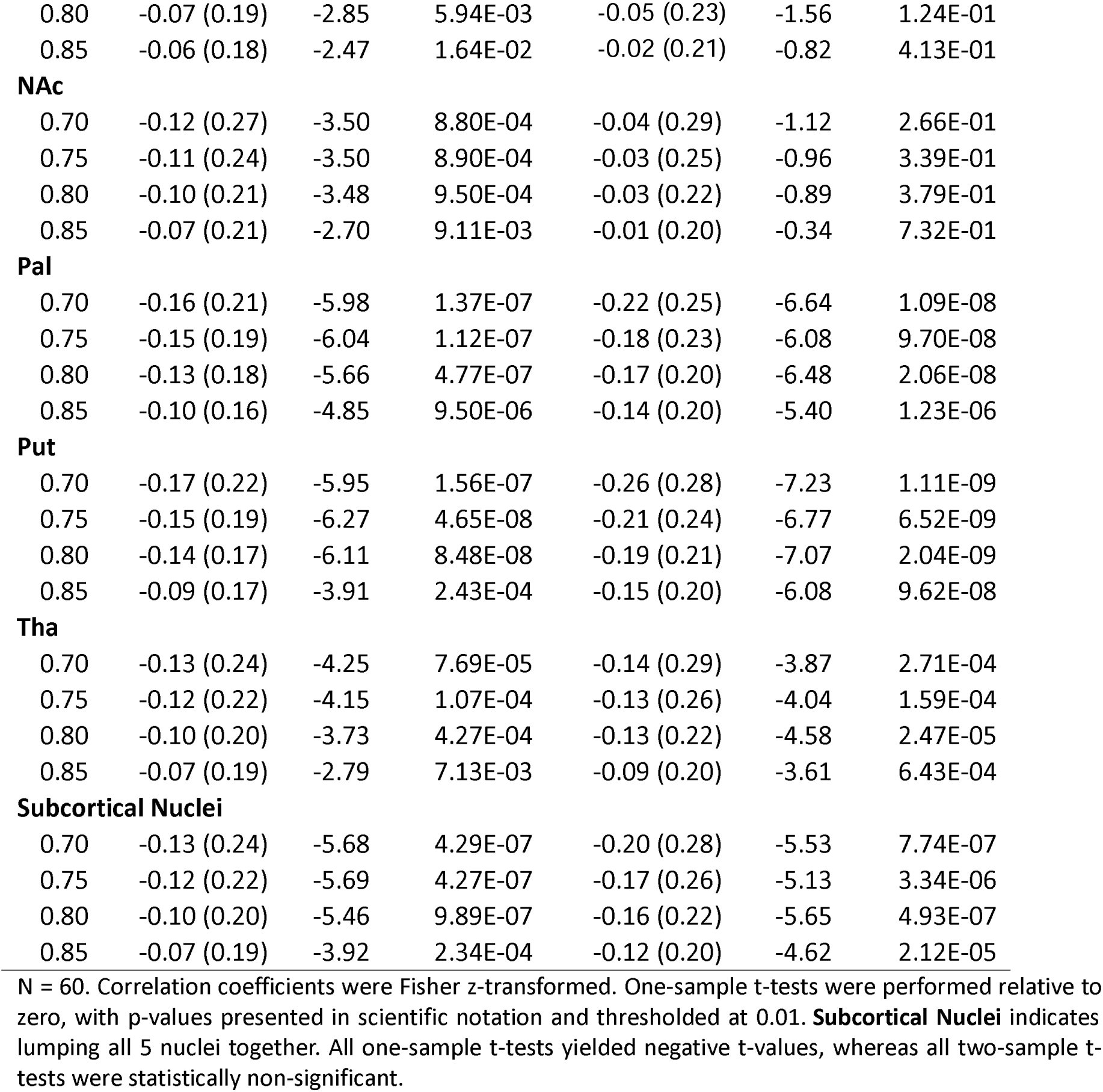
Correlation between frontal ALFF and fronto-subcortical connectivity (S2F)

|  | ADHD |  | one-sample t-tests |  | HC | one-sample t-tests |  |
| --- | --- | --- | --- | --- | --- | --- | --- |
| Gamma | mean (std) | t-scores | p-values |  | mean (std) | t-scores | p-values |
| **Cau** |  |  |  |  |  |  |  |
| 0.70 | -0.09 (0.24) | -2.77 | 7.49E-03 |  | -0.07 (0.31) | -1.77 | 8.26E-02 |
| 0.75 | -0.08 (0.21) | -2.83 | 6.41E-03 |  | -0.05 (0.27) | -1.50 | 1.40E-01 |

|  |  |  |  |  |  |  |
| --- | --- | --- | --- | --- | --- | --- |
| 0.80 | -0.07 (0.19) | -2.85 | 5.94E-03 | -0.05 (0.23) | -1.56 | 1.24E-01 |
| 0.85 | -0.06 (0.18) | -2.47 | 1.64E-02 | -0.02 (0.21) | -0.82 | 4.13E-01 |
| **NAc** |  |  |  |  |  |  |
| 0.70 | -0.12 (0.27) | -3.50 | 8.80E-04 | -0.04 (0.29) | -1.12 | 2.66E-01 |
| 0.75 | -0.11 (0.24) | -3.50 | 8.90E-04 | -0.03 (0.25) | -0.96 | 3.39E-01 |
| 0.80 | -0.10 (0.21) | -3.48 | 9.50E-04 | -0.03 (0.22) | -0.89 | 3.79E-01 |
| 0.85 | -0.07 (0.21) | -2.70 | 9.11E-03 | -0.01 (0.20) | -0.34 | 7.32E-01 |
| **Pal** |  |  |  |  |  |  |
| 0.70 | -0.16 (0.21) | -5.98 | 1.37E-07 | -0.22 (0.25) | -6.64 | 1.09E-08 |
| 0.75 | -0.15 (0.19) | -6.04 | 1.12E-07 | -0.18 (0.23) | -6.08 | 9.70E-08 |
| 0.80 | -0.13 (0.18) | -5.66 | 4.77E-07 | -0.17 (0.20) | -6.48 | 2.06E-08 |
| 0.85 | -0.10 (0.16) | -4.85 | 9.50E-06 | -0.14 (0.20) | -5.40 | 1.23E-06 |
| **Put** |  |  |  |  |  |  |
| 0.70 | -0.17 (0.22) | -5.95 | 1.56E-07 | -0.26 (0.28) | -7.23 | 1.11E-09 |
| 0.75 | -0.15 (0.19) | -6.27 | 4.65E-08 | -0.21 (0.24) | -6.77 | 6.52E-09 |
| 0.80 | -0.14 (0.17) | -6.11 | 8.48E-08 | -0.19 (0.21) | -7.07 | 2.04E-09 |
| 0.85 | -0.09 (0.17) | -3.91 | 2.43E-04 | -0.15 (0.20) | -6.08 | 9.62E-08 |
| **Tha** |  |  |  |  |  |  |
| 0.70 | -0.13 (0.24) | -4.25 | 7.69E-05 | -0.14 (0.29) | -3.87 | 2.71E-04 |
| 0.75 | -0.12 (0.22) | -4.15 | 1.07E-04 | -0.13 (0.26) | -4.04 | 1.59E-04 |
| 0.80 | -0.10 (0.20) | -3.73 | 4.27E-04 | -0.13 (0.22) | -4.58 | 2.47E-05 |
| 0.85 | -0.07 (0.19) | -2.79 | 7.13E-03 | -0.09 (0.20) | -3.61 | 6.43E-04 |
| **Subcortical Nuclei** |  |  |  |  |  |  |
| 0.70 | -0.13 (0.24) | -5.68 | 4.29E-07 | -0.20 (0.28) | -5.53 | 7.74E-07 |
| 0.75 | -0.12 (0.22) | -5.69 | 4.27E-07 | -0.17 (0.26) | -5.13 | 3.34E-06 |
| 0.80 | -0.10 (0.20) | -5.46 | 9.89E-07 | -0.16 (0.22) | -5.65 | 4.93E-07 |
| 0.85 | -0.07 (0.19) | -3.92 | 2.34E-04 | -0.12 (0.20) | -4.62 | 2.12E-05 |

1 N = 60. Correlation coefficients were Fisher z-transformed. One-sample t-tests were performed relative to 2 zero, with p-values presented in scientific notation and thresholded at 0.01. **Subcortical Nuclei** indicates 3 lumping all 5 nuclei together. All one-sample t-tests yielded negative t-values, whereas all two-sample t- 4 tests were statistically non-significant.

**Table 3.**
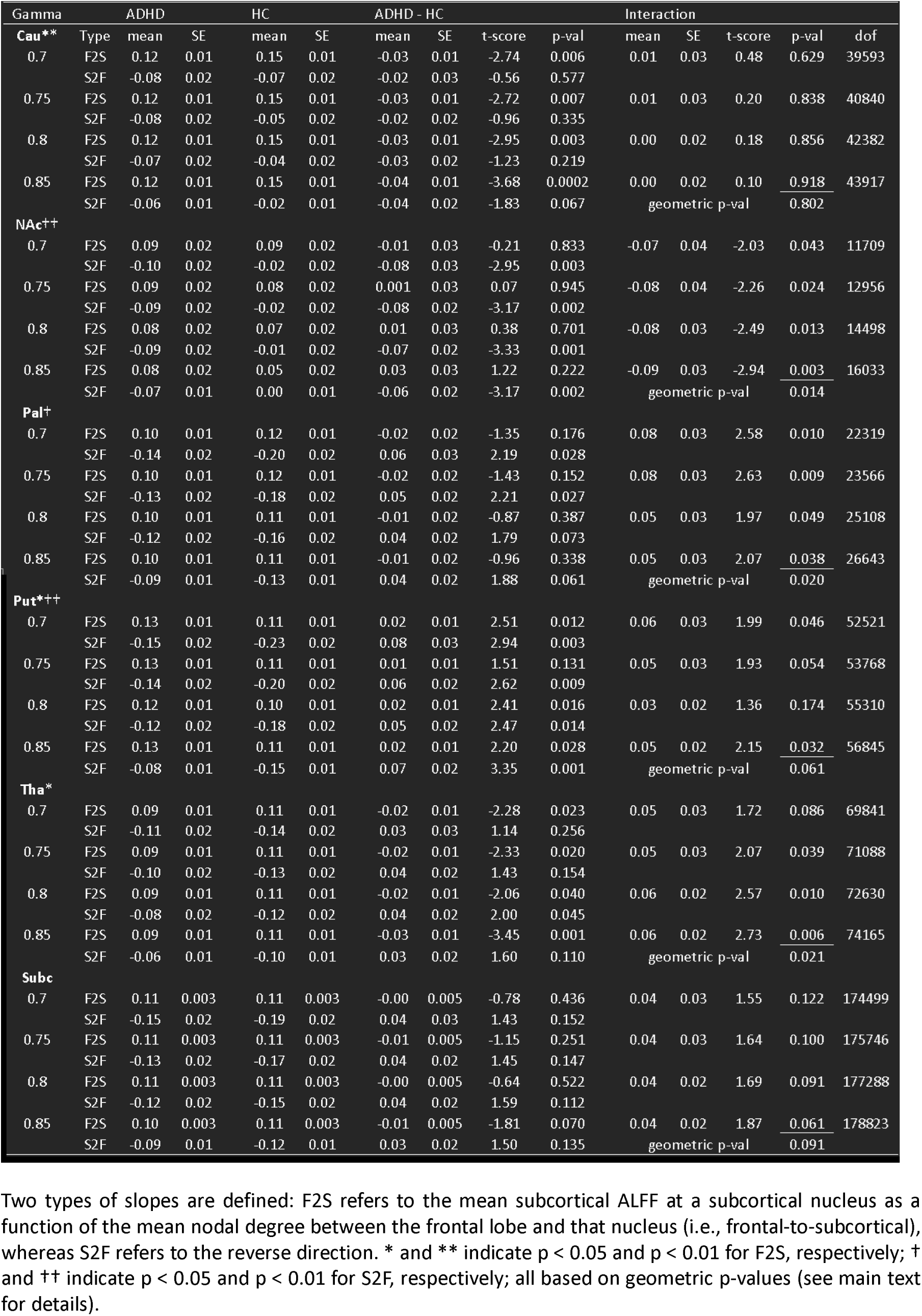
Results of linear mixed-effects (LME) analysis without random slope terms: estimated group-wise mean slopes, group contrasts, and interaction effects.

The strikingly opposite trends—indicative of frontal-to-striatal excitation and striatal-to-frontal inhibition—are shown in Figure 3. For illustration, the voxels from the five subcortical nuclei were aggregated to emphasize the contrasting patterns. The subplots in the left column contain more data points than those in the right column, as each voxel within the subcortical nuclei was treated as an individual analytic unit, whereas in the frontal lobe, the analytic units were defined based on MOSI-derived outputs. Consistent with prior findings on frontal–limbic interactions, the present results revealed asymmetrical slopes between nodal power and nodal degree, with stronger top-down modulation originating from the frontal lobe—i.e., F2S slopes were greater than S2F slopes—suggesting more prominent regulatory influence exerted by the FCX (Lee, 2025b).

**Figure 3.**
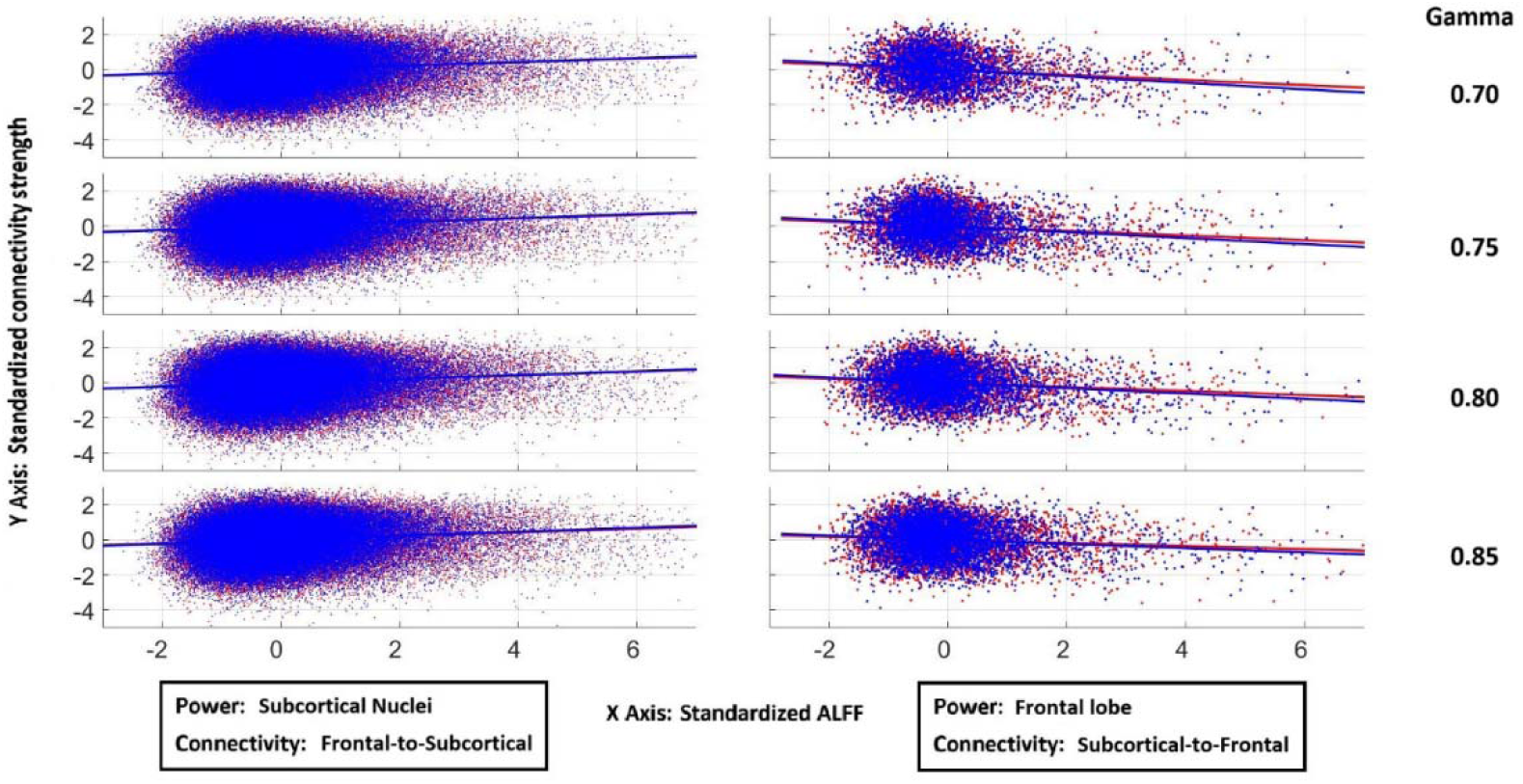
Scatter plots with regression lines illustrating the relationships between regional power and functional connectivity. Left column: voxel-wise ALFF within subcortical nuclei plotted against mean frontal-to-subcortical nodal strength. Right column: mean ALFF within the frontal lobe plotted against mean subcortical-to-frontal nodal strength. Rows correspond to four increasing Gamma values (0.70 to 0.85). The left and right columns show robust but opposing trends, as reflected in the opposite directions of the regression slopes. Blue indicates HC, and red indicates ADHD.

Interestingly, the association between nodal ALFF and nodal strength in the Cau-to-Frontal and NAc-to-Frontal conditions reached statistical significance in the ADHD group but not in the HC group, suggesting a potential interaction effect in line with the hypothesis. The corresponding two-sample t-tests (not shown in Table 2) were as follows: For Cau2F at gamma values of 0.70, 0.75, 0.80, and 0.85, the t-scores (p-values) were -0.30 (0.766), -0.54 (0.592), -0.58 (0.561), and -1.00 (0.319), respectively; while For NAc2F at gamma values of 0.70, 0.75, 0.80, and 0.85, the t-scores (p-values) were -1.54 (0.126), -1.69 (0.094), -1.75 (0.082), and -1.70 (0.092), respectively. A potentially significant interaction effect was observed in the NAc, to be examined using LME models below.

### Analysis II—LME analyses between ALFF and nodal strength

In the LME analysis, the geometric mean p-values for the F2S and S2F conditions across 4 gamma values of the Cau, NAc, Pal, Put, and Tha were (0.002, 0.231), (0.592, 0.002), (0.243, 0.043), (0.029, 0.004), and (0.012, 0.118), respectively (Table 3). An interaction effect in NAc approached significance (p = 0.014). Notably, under a more complex—and arguably overly stringent—LME model incorporating random slopes, all main and interaction effects in subcortical regions were negative except for the significant interaction effect in NAc, as shown in Table 4 and Figure 4 (geometric p = 0.009; additional details in Table S4, **Supplementary Materials**). There appear to be four distinct patterns in the main effects (ADHD minus HC): (1) less positive in F2S and less negative in S2F—Tha, Pal, and the combined subcortical nuclei; (2) less positive in F2S— Cau; (3) more negative in S2F—NAc; and (4) more positive in F2S and less negative in S2F—Put. The implications to the neuropathology of ADHD are elaborated in the **Discussion**.

**Figure 4.**
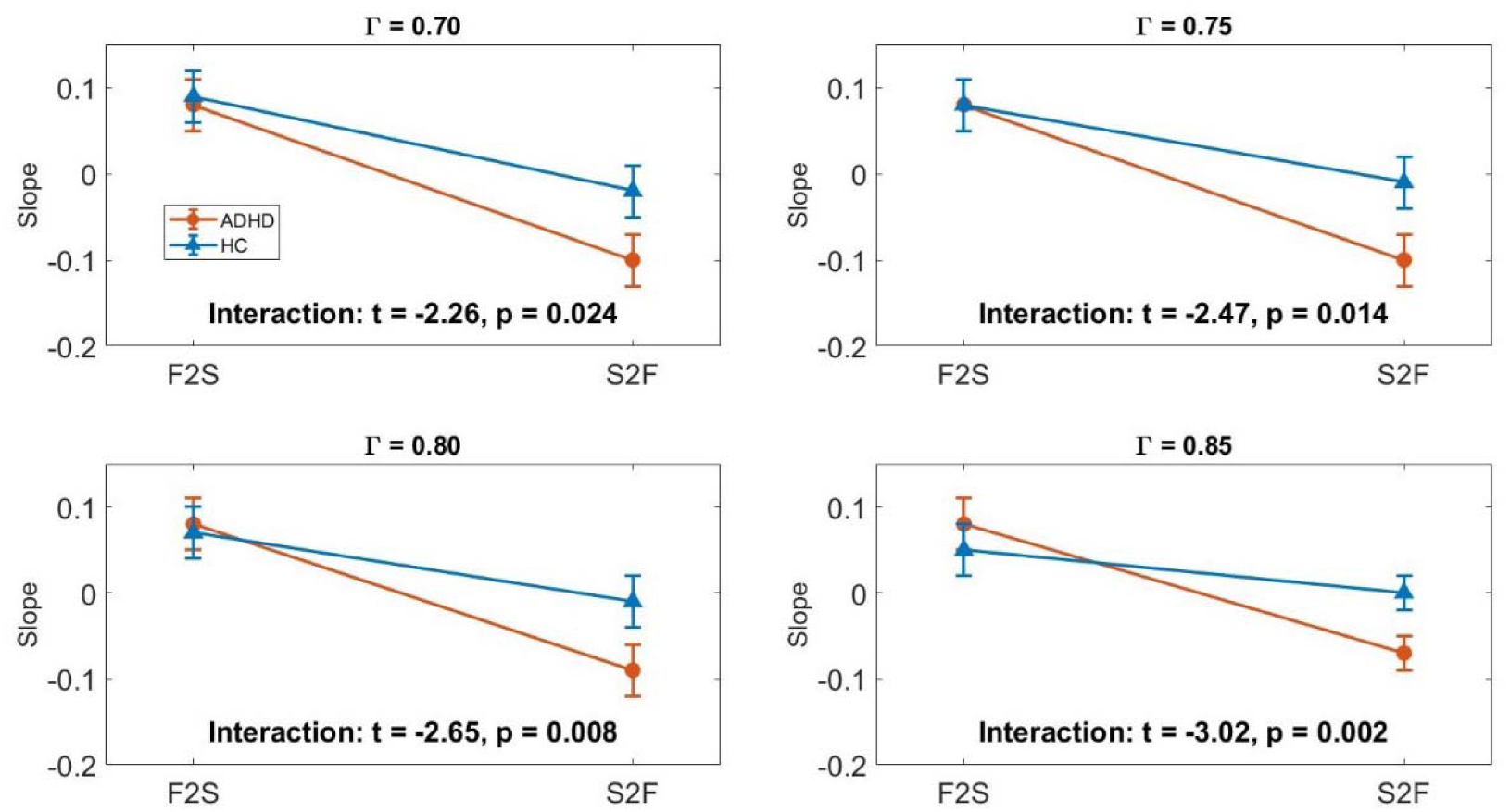
Relationships between ALFF and nodal strength in the NAc and frontal lobe based on slopes derived from Linear Mixed Effects (LME) models at four Gamma values. F2S: slopes of ALFF at NAc against mean functional connectivity between NAc and frontal lobe. S2F: slopes of mean ALFF at frontal lobe against mean functional connectivity between NAc and frontal lobe. Data are shown for two groups, ADHD (triangles) and HC (circles), with error bars representing standard errors. Interaction effects between group and condition are reported below each subplot.

**Table 4.**
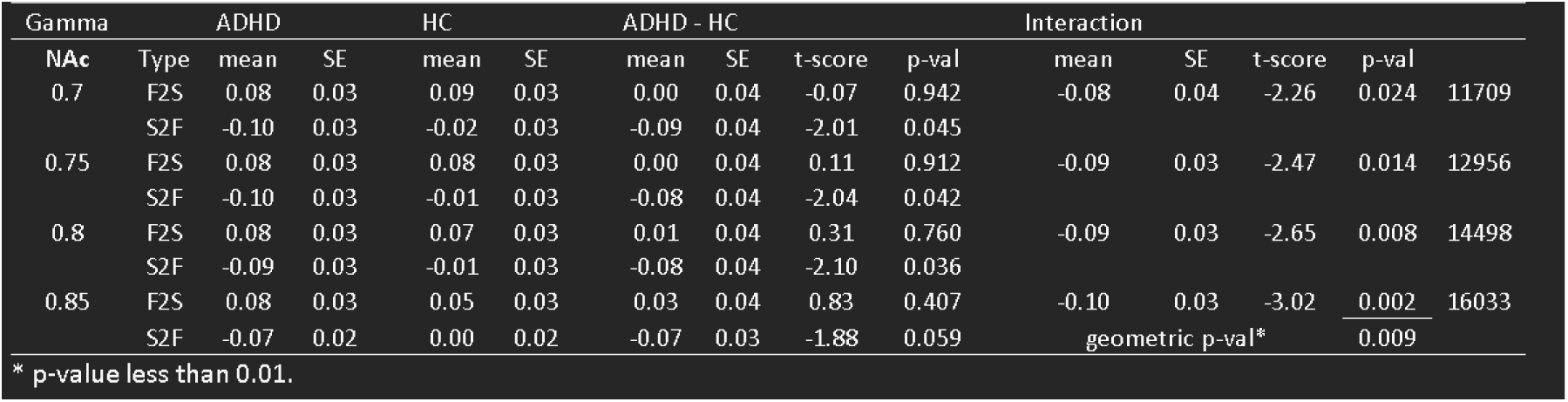
Results of linear mixed-effects (LME) analysis with random slpoe terms: estimated group-wise mean slopes, group conrasts, and interaction effects (NAc subset of full model)

For completeness, between-group comparisons of ALFF and mean nodal strength were conducted for the five selected subcortical nuclei (either functionally connected to the frontal lobe or within subcortical regions) as well as within the frontal lobe. Except for a trend toward higher nodal strength at the NAc in the ADHD group within the subcortical network (p = 0.097, t = 1.67), all other comparisons yielded non-significant results. Detailed findings are presented in Parts II and III of the **Supplementary Materials**.

## Discussion

This study aimed to empirically evaluate a neuropathological model of ADHD grounded in dialectic neuroscience (Lee, 2025a). The model contends that aberrant descending fronto-subcortical projections may constitute a core developmental vulnerability, impacting both ventral (predominant in ADHD) and dorsal fronto-striatal pathways. Within the ventral circuitry, dysfunction in the autoregulatory FCX–VTA loop is posited as the primary locus of disruption in ADHD, with downstream propagation to the NAc further contributing to atypical reward processing and limbic dysregulation. The novel approach developed in this study enabled directional characterization of fronto-striatal interactions, revealing a consistent dissociation across subcortical targets and MOSI resolutions—excitatory in the F2S direction and inhibitory in the S2F direction. A steeper positive slope in the F2S condition suggests that the frontal lobe may more effectively excite subcortical nuclei, likely via glutamatergic transmission (Groenewegen and Trimble, 2007), whereas a steeper negative slope in the S2F condition indicates that subcortical nuclei may impose stronger inhibitory influences on the FCX. These opposing slopes in the F2S and S2F conditions may provide a valuable framework for examining the network balance between frontal and subcortical systems. The group comparison results were broadly consistent with the model’s predictions. First, in the F2S condition, correlations between nodal strength and power were lower in ADHD than HC across several subcortical structures, including the Cau, Pal, and Tha, as well as when all subcortical nuclei were considered collectively (Subc). Second, the NAc demonstrated a robust interaction effect, replicated across two independent LME models. The observed pattern of reduced frontal volume and increased NAc-centered functional connectivity aligns with prior reports (Norman et al., 2016, Norman et al., 2024).

Overall, group comparisons revealed parallel shifts in both F2S and S2F slopes within Pal, Tha, and Subc—specifically, reduced F2S positivity and attenuated S2F negativity—suggesting a preserved excitation–inhibition balance in these nuclei in ADHD. Notably, the Tha and Subc patterns further supports the view that, when considering the common relay station of striato-thalamo-cortical circuitry and the subcortical nuclei collectively, the overall excitation–inhibition balance remains intact. A similarly reduced F2S slope was also observed in Cau. The consistent reduction in F2S across Pal, Tha, Cau, and Subc lends support to the first prediction. However, this balance appears disrupted in the motor-related relay station Put, which exhibits increased F2Put and decreased Put2F in ADHD. This shift—marked by heightened excitation relative to inhibition— may lead to maladaptive activation of the motor system, primarily contributing to the high comorbidity with tic disorders, but potentially also to hyperactivity (Wanderer et al., 2021, Rothenberger and Heinrich, 2022), although the precise neural mechanisms remain to be clarified.

Among the subcortical nuclei, the caudate and NAc are most closely implicated in ADHD (Lee, 2025a). In HC, the slope mediating nodal degree versus nodal power of S2F at Cau (Cau2F) did not reach significance, contrasting with motor-related nuclei such as Put and Pal, which showed balanced F2S excitation and S2F inhibition. Consistent with the findings, fiber tracking research of the thalamus exhibits distinct patterns of reciprocal connectivity across subregions related to cognitive and motor basal ganglia loops. Specifically, the mediodorsal thalamic nucleus, primarily connected with Cau and prefrontal cognitive (and affective) circuits, shows substantial nonreciprocal corticothalamic projections from diverse frontal cortical areas. In contrast, the ventral anterior and ventrolateral thalamic nuclei, associated with Put and Pal within motor circuits (including the supplementary motor area, motor and premotor cortices), exhibit stronger reciprocal thalamocortical and corticothalamic connections. This differential organization aligns with the findings, as motor loops maintain tightly coupled, bidirectional thalamocortical communication, whereas cognitive loops display broader, asymmetric cortical inputs to thalamic nuclei, reflecting their integrative, associative and regulatory roles. These distinctions likely underlie fundamental differences in excitation-inhibition balance demands between cognitive and motor basal ganglia-thalamocortical systems (Grahn et al., 2008, McFarland and Haber, 2002). Interestingly, neither the slope of NAc2F nor F2NAc reached significance in the HC group, which is understandable given the diffuse and complex afferent and efferent connections of the NAc. These involve limbic and deep subcortical structures beyond the cortico-striato-thalamo-cortical circuits (Alexander et al., 1986), thereby diluting its relationship with the frontal lobe (Sesack and Grace, 2010, Groenewegen and Trimble, 2007).

However, in the ADHD group, the slope of NAc2F was significantly more negative than in HC, making it the only subcortical nucleus to show a significant interaction effect—thus supporting the second prediction. In addition to VTA-FCX, the NAc also plays a central role in the neuropathology of ADHD (Lee, 2025a), becoming sensitized and disinhibited due to low tonic DA, which reduces presynaptic D2 receptor–mediated inhibition, and to hypofrontality, which weakens GABAergic regulation from the VTA (Carr and Sesack, 2000, Goto and Grace, 2005). This low tonic DA state may, in turn, trigger compensatory enhancement of phasic DA signaling (Grace, 1991, Grace, 1995). Together, these mechanisms result in hyperactivity of the NAc, and consequently, the mesolimbic system. Through reciprocal suppressive interactions between the limbic system and the FCX (Lee and Xue, 2018, Lee, 2025b), hyperactivity in the NAc may ultimately exert strong inhibitory influences on the FCX in ADHD—a pattern reflected in the analytic results. A similar, though non-significant, enhancement of S2F suppression in the caudate observed in the ADHD group may be driven by shared underlying hypo-dopaminergic states (Lee, 2025a). Hypoactivity in the FCX can reduce DA levels in the Cau (Strafella et al., 2001, Chéramy et al., 1986), and hypodopaminergism is a key contributor to hypofrontality in ADHD. An elevated Cau2F nodal degree may indicate a shared dopaminergic deficiency between specific module(s) in the FCX and the Cau, which could further aggravate hypofrontality and manifest as increased Cau2F inhibition. In contrast, FCX modules less involved in caudate DA metabolism may exhibit weaker correlations and consequently show higher ALFF.

## Conclusion

This study empirically tested a neuropathology model of ADHD derived from dialectic neuroscience, which posits that aberrant descending fronto-subcortical projections— particularly those involving the FCX–VTA loop—constitute a core developmental vulnerability. Leveraging a novel directional framework, a consistent dissociation between F2S excitation and S2F inhibition across subcortical targets was identified, providing a foundation for examining directional network properties in ADHD. Group comparisons supported the model: diminished frontal drive (lower F2S correlations) was evident across multiple subcortical nuclei, and a robust interaction centered on the NAc aligned with prior evidence of mesolimbic dysregulation. More broadly, this work exemplifies the utility of dialectic neuroscience—a theory-driven, mechanistically anchored alternative to traditional psychological paradigms and data-driven approaches—as a promising direction for understanding psychiatric disorders at the systems level.

## Data Availability

All data produced are available online at https://fcon_1000.projects.nitrc.org/indi/adhd200/

